# Thoughts on Higher Medical Education Under Major Public Health Emergencies: Thinking Ahead After COVID-19 Outbreak

**DOI:** 10.1101/2020.04.06.20053918

**Authors:** Wei Lin, Yan Chen, Songchang Shi, Jixing Liang, Huibin Huang, Liantao Li, Liangchun Cai, Liyao Zong, Nengying Wang, Junping Wen, Gang Chen

## Abstract

**Importance:** The spread of coronavirus disease 2019 (COVID-19) has posed great threat to people’s health and several medical schools in the world suspended classes as a precaution against the virus. China has also adopted precautionary measures to keep medical schools running without suspending classes. Thinking ahead after COVID-19 Outbreak is important.

**Objective:** To explore the most suitable teaching and learning pattern in medical school during COVID-19 Outbreak.

**Design:** This study is a case-control study. We had tried to apply a new blended teaching model based on 5G network that combined team-based learning (TBL) and online interaction to the students before the outbreak and then universities responded to the COVID-19 outbreak by closing campuses and shifting to other forms of distance learning. In other word, the courses started using blended teaching model before COVID-19 outbreak and might last using other forms of distance learning throughout the pandemic. Five Point Likert Scale Questionnaires which contains 20 items were used, and the effect of the two kinds of teaching patterns was compared by evaluating the indicators of core competencies of students including professionalism, attitude towards learning, knowledge and learning skills, teamwork skills, motivation in learning, adaptability and acceptance of the courses and network environment.

**Setting:** Our study based on a single center.

**Participants:** Fifty fourth-year medical students receiving the “5+3” pattern courses regarding internal medicine were enrolled in the study.

**Exposure(s) (for observational studies):** The teaching and learning patter started using blended teaching model before COVID-19 outbreak and might last using other forms of distance learning throughout the pandemic.

**Main Outcome(s):** According to the descriptive statistical analysis of the first part of the questionnaire (question 1-16), the average score of adaptability and acceptance of the courses is 2.60 lower than 3, indicating that students are more adapted to other forms of distance learning during COVID-19 outbreak; the average score of the rest of the questions is higher than 3, indicating that blended teaching model based on 5G network is superior to other forms of distance learning. The number of male students who are inclined to the blended teaching model based on 5G network is 0.13 times as much as that of female students (95%CI:0.028∼0.602, p=0.009).

**Results:** Online forms of distance learning were accepted by the students. Female students had higher expectations on the course and were more likely to adapt well to the change during the COVID-19 outbreak. However, all students preferred the blended teaching model based on 5G network that combined team-based learning (TBL) and online interaction before the pandemic.

**Conclusion:** It indicates that medical education based on 5G network that combined team-based learning (TBL) and online interaction is a more suitable option to teach medical students online. China’s experience in online higher medical education may serve as a reference to other countries during the pandemic.

**Key point:** *Questions:* What are the reflections on approaches to teaching and learning during COVID-19 Outbreak?

*Findings:* Fifty fourth-year medical students receiving the “5+3” pattern courses regarding internal medicine were enrolled. Five Point Likert Scale Questionnaires which contains 20 items were used. This study indicates that medical education based on 5G network that combined team-based learning (TBL) and online interaction is a more suitable option to teach medical students online during COVID-19 outbreak.

*Meaning:* China’s experience in online higher medical education may serve as a reference to other countries during the pandemic.

## Introduction

The advances in medical education is an important indicator of a country’s level of medical science development. The core mission of medical education is to train qualified medical personnel for the country. In today’s era of globalization, medical education associated with human health issues such as the control of HIV/AIDS, the outbreak of SARS and the spread of bird flu are no longer limited by national borders.

Coronavirus Disease 2019 (COVID-19) has been ongoing in China since December, 2019. On January 30, 2020, the second meeting of the Emergency Committee convened by the WHO Director-General under the International Health Regulations (IHR) declared that the outbreak of COVID-19 constitutes a Public Health Emergency of International Concern (PHEIC)^[1]^. The pandemic raised great concern in China and other countries [2-5]. In order to prevent the spread of COVID-19 at mass gatherings, Ministry of Education of the People’s Republic of China adopted countermeasures such as online education to keep medical schools running without suspending classes. According to Coronavirus disease 2019 (COVID-19) Situation Report as of 18 March 2020, the total confirmed cases globally reached 191127 with 110011 confirmed cases outside China including 15084 total confirmed new cases with an increase of over 13%. This pandemic has posed great challenges to the global public health, health care system and medical education, which triggered new thinking about medical education under PHEIC. This article summarizes and reflects on the measures adopted by China in medical education during COVID-19 outbreak so as to provide vital experience of how to conduct medical education under PHEIC.

*Health professionals for a new century: transforming education to strengthen health systems in an interdependent world* published in Lancet proposed an idea of the third-generation educational reform^[6]^. It focuses on systems based medical education to improve the performance of health systems by adapting core professional competencies to specific contexts, while drawing on global knowledge. With the rapid development of medical science, the traditional teaching methods featuring slow knowledge updating and low teaching efficiency, to some extent, are unable to fully meet the diverse learning needs of medical students. Since blended learning was proposed in 2003, it has become one of the mainstream educational approaches in the medical educational reform^[7]^. Blended learning is a style of education in which students learn via electronic and online media as well as traditional face-to-face teaching, which places an emphasis on empowering students with the skills and knowledge required to make the most of the online material and independent study time, guiding students toward the most meaningful experience possible^[8]^. E-learning is based on network support. 5G is the abbreviation of the fifth-generation mobile communication technology. 5G network features enhanced mobile broadband (eMBB) coping with the significant increase of data volumes, overall data capacity, and user density, massive machine type communications (mMTC) requiring low power consumption for a huge number of connected devices, ultra-reliable and low latency communications (URLLC) providing safety-critical and mission critical applications.

Compared with 4G network, WLANs or Wi-Fi, 5G network guarantees the same consistency in performance with less interference, which is very important for critical machine-to-machine communications, especially where the number of devices is massive. Team-based learning (TBL) is a structured form of small-group learning that emphasizes student preparation out of class and application of knowledge in class. In accordance with boosting medical educational reform by new technologies, we had tried to apply a new blended teaching model based on 5G network that combined TBL and online interaction to 50 fourth-year medical students who were receiving the “5+3” pattern courses regarding internal medicine in July, 2019 before the outbreak of COVID-19. They were organized strategically into 5 teams of 10 students that work together throughout the online class in this model. The pandemic was later determined to be PHEIC seriously endangering international public health, which posed great challenges to medical education. Medical universities in China closed campuses and shifted to other forms of distance learning such as massive open online courses (MOOCs), recorded teaching video in Rain Classroom, chatting via QQ in response to the COVID-19 outbreak. Home-based learning allows students to extend their education from school to home, from offline to online, from passive learning to active learning. It seems to be an alternative learning approach under PHEIC that conform to the trend of the third-generation educational reform. In this study, we compared the blended teaching model based on 5G network that combined TBL and online interaction conducted on July, 2019 before the outbreak and other forms of distance learning during the pandemic by using online survey tool to discuss the impact of this change will bring towards the students’ core competencies including professionalism, attitude towards learning, knowledge and learning skills, teamwork skills, motivation in learning, adaptability and acceptance of the courses.

## 2. Subject and Method

### 2.1 Subject

Fifty fourth-year medical students receiving the “5+3” pattern courses regarding internal medicine were enrolled in the study. “5+3” pattern is an innovative training pattern in graduate education for a professional degree in clinical medicine. The pattern focuses on innovation in training talent as well as reform of the administrative mechanism. In “5+3” pattern, medical students directly enter the 3 years of pursuing a master ‘s degree after completing 5 years of preclinical and clinical courses. It advances the organic link between student cultivation and standardized training for residents, and facilitates the tight connection between professional training and occupational qualifications. The study protocol was approved by the Ethics Committee of Fujian Provincial Hospital, and all the participants provided written informed consent.

### 2.2 Study design

The questionnaire in the electronic form was distributed to the students via WeChat. They filled it out voluntarily and submitted the statistics the next day. It was designed through reviewing domestic and foreign literature such as Health professionals for a new century: transforming education to strengthen health systems in an interdependent world published in 2010 and the “Six Excellence and One Top-Notch Talent Training Plan 2.0” launched by Ministry of Education of the People’s Republic of China in 2018.

The Five Point Likert Scale Questionnaire regarding the two kinds of teaching patterns which contains 20 items was used to compare the blended teaching model based on 5G network that combined TBL and online interaction and other forms of distance learning. It consists of 5 answer options which contain two extreme poles and a neutral option connected with intermediate answer options. The questionnaire was divided into two parts. The first part refers to the comparison between the effect of two kinds of teaching patterns. The options start with “strongly in favor of other forms of distance learning “at 1 point, “neutral” at 3 point, and “strongly in favor of the blended teaching model based on 5G network that combined TBL and online interaction “at 5. The second part refers to the comparison between the user experience of online platform based on 5G network and other prevalent forms of wireless internet connection such as WIFI and 4G network. The options start with “strongly in favor of the user experience of other prevalent forms of wireless internet connection such as WIFI and 4G network “at 1 point, “neutral” at 3 point, and “strongly in favor of the user experience of online platform based on 5G network “at 5.

### 2.3 Statistical analysis

All data were collected by WJX, an electronic questionnaire design tool. They were verified by two teachers and exported to SPSS version 26.0 (IBM, United States). Reliability analysis and descriptive statistical analysis was conducted on the data, respectively. If data were normally distributed, they were expressed as mean ± standard deviation. If data were not normally distributed, they were expressed as the median. In addition, the binary logistic regression was used to analyze the impact of gender on the scores of the questionnaire.

## 3 Result

### 3.1 Basic information of the subjects

Fifty fourth-year medical students receiving the “5+3” pattern courses regarding internal medicine were enrolled in the study. Fifty electronic questionnaires were distributed and 96% (48/50) of the valid ones were returned. The subjects aged (22.4 ± 0.79) years old, of which 25 were male students, accounting for 52.0% of the effective sample; 23 were female students, accounting for 48.0% of the effective sample.

### 3.2 Data analysis

#### 3.2.1 Reliability analysis

Cronbach’s Alpha in SPSS Statistics was used to measure the internal consistency of the valid questionnaires in this study. The results showed that the questionnaire has high internal consistency or high reliability as the value of Cronbach’s Alpha is 0.73, and the value of Cronbach’s Alpha based on the standardized items is 0.95, both of which are at a high level.

#### 3.2.2 Descriptive statistical analysis

The questionnaire in this study consists of two parts involving five aspects of indicators of core competencies (Figure 1). The first part of the questionnaire is designed on the basis of competency-oriented teaching. The items cover four indicators of core competencies including A:professionalism and attitude towards learning (question 1-5), B: knowledge and learning skills (question 8,9,10,12,16), C: teamwork spirit (question 11), as well as D: motivation in learning and acceptance of the courses (question 6,7,13,14,15). The second part of questionnaire is mainly for the evaluation of network environment of online education including E: user experience under the certain network environment (question 17-20).

**Figure 1.**
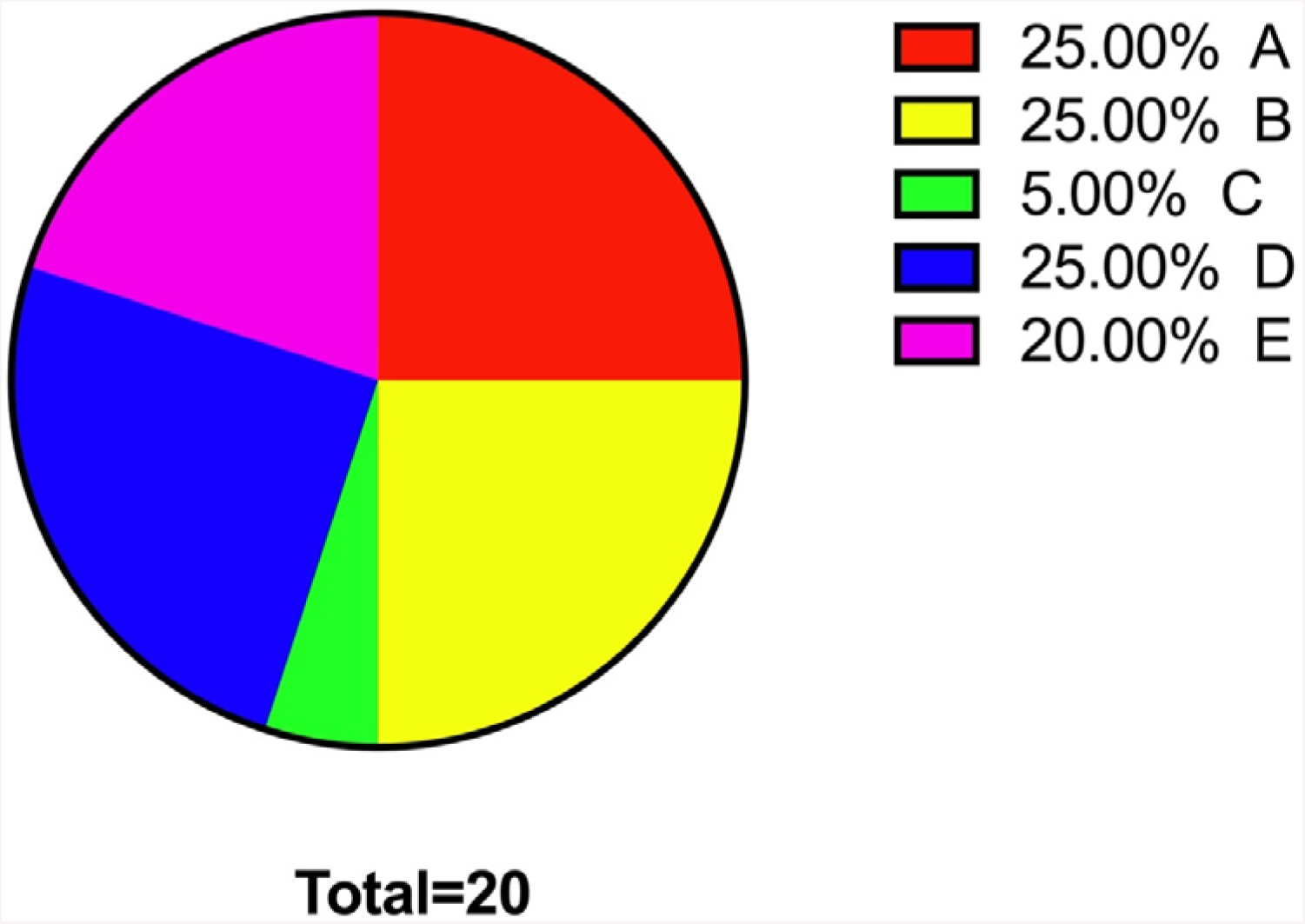
The composition of indicators of core competencies in the questionnaire. A: Professionalism and attitude towards learning (question 1-5) B: Knowledge and learning skills (question 8,9,10,12,16) C: Teamwork spirit (question 11) D: Motivation in learning and acceptance of the courses (question 6,7,13,14,15) E: User experience under the certain network environment (question 17-20).

The data of the results of the questionnaire were normally distributed(P> 0.05). According to the descriptive statistical analysis of the first part of the questionnaire (question 1-16) (Table 1), the average score of adaptability and acceptance of the courses is 2.60 lower than 3, indicating that students are more adapted to other forms of distance learning during COVID-19 outbreak; the average score of the rest of the questions is higher than 3, indicating that blended teaching model based on 5G network is superior to other forms of distance learning on the aspects of professionalism, attitude towards learning, knowledge and learning skills, teamwork skills and motivation in learning; the average score of the challenging level of the course is 4.04, indicating that blended teaching model based on 5G network is more challenging for most of the students than other forms of distance learning.

According to the descriptive statistical analysis of the second part of the questionnaire (question 17-20) (Table 2), the average score of network latency and stability is 2.83 lower than 3, which suggests that other forms of wireless internet connection are more stable currently; the rest of the questions in the second part of the questionnaire score more than 3, indicating that students believe that blended teaching model based on 5G network is better than other forms of distance learning on the aspects of real-time interaction, high-definition image quality and voice clarity.

#### 3.2.3 Binary logistic regression

Expectations on the course will have an impact on students’ professionalism, attitude towards learning (question 1, Table 3). As for this question, it shows that the number of male students who are inclined to the blended teaching model based on 5G network is 0.13 times as much as that of female students (95%CI:0.028∼0.602, p=0.009). In other word, female students have higher expectations on the blended teaching model based on 5G network. Moreover, the impact of the course on the comprehensive abilities and advanced thinking are highly related to the knowledge and learning skills which is one of the core competencies (question 10, Table3). It shows that the number of male students who are inclined to the blended teaching model based on 5G network is only 0.163 times as much as that of female students, which indicates that female students believe the blended teaching model based on 5G network is beneficial to improving their comprehensive abilities and advanced thinking. As for question 17-20, it shows that there is no difference between male and female in terms of online platform preferences,

## 4 Discussion

### 4.1 Importance and necessity of the strategies for higher medical education in China during PHEIC

Before the COVID-19 pandemic, China’s higher medical education mostly adopted traditional education model, namely the second-generation education model. However, with the incredible growth of Internet since the 1990s, online education has begun to thrive. In 2009, Khan Academy and flipped class became popular in China, and a number of higher medical education institutions in China have begun to accept or try online education. In 2010, large-scale online courses such as MOOCs had a tremendous impact on China’s higher medical education. At present, there are numerous excellent online platforms of MOOCs, including Coursera, Udacity and edX, the top three world-renowned education platforms as well as domestic platforms such as Chinese university MOOC, XuetangX and Icourses. The proliferation of online education has already been an inevitable trend, and the COVID-19 pandemic facilitates its development.

There have also been some special periods in Chinese medical education history that are very similar to the current COVID-19 pandemic. For example, during the outbreak of SARS in 2003, classes were suspended in Beijing and some other cities in China. At that time the Internet was not as developed as it is now, so educational programs were mostly broadcasted on air to guide students to learn independently.

These measures played an important part in special times, and also accelerated the rapid development of education informatization and the infrastructure. As speculated earlier, the findings of this study show that medical students prefer home-based online learning in terms of adaptability and acceptance of the course. With regard to the relevant observational indicators of competency-oriented medical education involved in this research questionnaire, the results also show that quite a number of medical students believe that home-based online education model can, to some extent, cultivate their professionalism, stimulate their motivation in learning and bring more fun to the study amid the pandemic. Meanwhile, medical students can obtain knowledge and improve learning skills through online education. In a sense, through online interaction based on WeChat groups, QQ groups and other platforms, home-based online learning can also foster their teamwork spirit. Therefore, the result of this study to some degree further confirm that Chinese government has implemented timely and correct strategies in response to the PHEIC, which extends higher medical education from school to home and accelerated the popularization of online education. It may be an inevitable trend for the development of medical education in the future.

### 4.2 The online teaching model of higher medical education in China under PHEIC

After the COVID-19 outbreak, students had to stay at home to eliminate chances of being infected. However, the devices and network environment for online learning differ among families. Given the Guidance on Online Education issued for schools in China amid COVID-19 pandemic, China’s higher medical education was conducted via PCs, mobile devices, TV and other media based on online platforms. The teaching models include online courses, live streaming, independent learning, etc.

Online courses are a series of lessons delivered on the website or to the mobile devices, which can be conveniently accessed anytime and anyplace. In recent years, with the booming of high-quality course resources, MOOCs and “golden courses” in the field of higher education and the popularity of micro courses and flipped classroom, the number of various online courses in high quality is increasing rapidly. In the face of COVID-19 pandemic, such massive resources can be accessed for free on the online platforms in China such as China University MOOC, Good University Online, Wisdom Tree, etc. As for the current teaching plan, it is recommended to employ MOOCs and SPOCs, meanwhile setting up Q&A groups as supplement for online courses. However, some shortcomings of online courses surfaced in its current application. For instance, the number of high-quality MOOCs, micro courses and “golden courses” are limited, which cannot cover all courses for all grades. As for the Internal Medicine courses, there is a lack of sufficient high-quality online course resources. Meanwhile, considering the nature of online courses, teachers cannot monitor students’ learning efficiency and test their real-time learning effectiveness. Thus, the following related measures are needed to improve the quality of online courses. ⍰ Publish daily learning tasks at a fixed time to create a good online learning atmosphere, and check students’ learning tasks in a timely fashion. ⍰ Offer learning support services, such as designating an excellent student as an assistant, managing class discipline, and collecting questions raised by students. ⍰ Organize live streaming classes and answer students’ doubts to strengthen their sense of actually being in the class. ⍰ Drawing up a plan to promote high-quality MOOCs, “golden courses” and micro courses after the pandemic.

Live streaming teaching, as a new type of online teaching model, is becoming more familiar to and applied by increasing numbers of teachers and students. Compared with online courses, live streaming teaching allows teachers and students to communicate in real time, provides a strong sense of presence and offers more interactions. Especially with the development of internet and popularity of live streaming culture, it appeals to college students even more. Live streaming teaching is a teaching model that relatively resembles offline classes but unstable network environment like high network latency and low network speed may ruin the students’ experience of learning in the live streaming online classroom. Hence, followings are some advice to improve the quality of live streaming teaching : ⍰ Familiarize with the live streaming platform in advance and test it, and make precautionary plans for unstable network environment. ⍰ Promote the application of 5G network in live streaming teaching and increase the bandwidth. The results of this study show that image quality, voice clarity, network speed and latency of 5G network are all better than other forms of wireless network connection. ⍰ In the process of teaching, course content can be divided into several fragments or small units, and some content-related interactions and tests can be presupposed. ⍰ With flipped classroom, students can study on their own first, find some common problems through exercises, tests and tasks, and then attend the live streaming to solve those problems. ⍰The teacher ought to turn on the camera throughout the teaching process to maintain a sense of presence. ⍰ Avoid the online rush hours to ensure a stable live streaming during the pandemic.

Independent learning model mainly depends on individual student to study on their own with learning resources provided by teachers while teachers give guidance and answer questions online afterwards. Under the circumstance where teachers and students are in different places, such self-learning is almost inevitable with advantages like low cost, easy operation and high flexibility, but the disadvantages lie in its weak interaction, poor learning atmosphere, and high requirements for self-discipline. During the pandemic, it may become a preferential option for most schools, particularly for teachers who lack relevant experience in online teaching, or the network environment at students or teachers’ home is poor.

In summary, it is recommended teaching models such as online courses, live streaming and independent learning can be adopted amid COVID-19 pandemic to achieve a better study experience. The result of the questionnaire show that students respond well to online education and have high enthusiasm, especially female medical students who have higher expectations for online courses. The 5G network can present high-definition image quality, improve voice clarity, and make real-time interaction more appealing. However, in terms of observation indicators concerning the evaluation of competence-oriented capacity, most medical students are more inclined to use the blended teaching model based on 5G network that combined TBL and online interaction. That indicates online learning based on 5G network can better improve students’ enthusiasm and sense of participation. We can try to diversify the form of online education to enhance the students’ learning experience. In particular, the model of flipped classroom can be adopted. Students in this model are encouraged to study independently through high-quality MOOCs and literature before class, and TBL can be carried out on social platforms such as online WeChat group or QQ group to cultivate students’ teamwork spirit. After that, teachers summarize the class by integrating knowledge of medical humanity, public health and social economics to improve students’ professionalism and analyzing the problems or questions raised by the students in the form of online live broadcast supported by 5G network to achieve a good learning experience.

### 4.3 Some suggestions on online teaching for different practitioners of higher medical education in China under major public health emergencies

During the COVID-19 pandemic, a variety of online teaching programs aimed at higher medical education has been formulated and implemented, but in this particular period the effectiveness and results of online teaching need joint efforts by subjects of practice involved in actual work such as higher medical education managers, teachers and students. ⍰ Amid current pandemic, the administrative department of higher medical education and school administrators should take actions based on a long-term perspective, systematic thinking and top-level design. ⍰ Teachers are the main body of the action of “keep medical schools running without suspending classes”. On the one hand,teachers have to change their ideas to form a “student-centered” teaching concept. On the other hand, teachers ought to improve their information literacy, familiarize themselves with online education, and master the method of online teaching. In addition to teaching basic professional knowledge, ideological and political concept, benevolence to patients, contribution to society, as well as the preventive medicine should be integrated into online courses. Moreover, it is necessary to raise medical students awareness of PHEIC during the pandemic, especially teach them about precautionary and preventive measures such as clinical diagnosis and treatment, isolation of patients and self-protection so that they can be competent in the future to handle public health emergencies and give medical guidance to the masses.

## 5. Conclusion

“Keep medical schools running without suspending classes” is China’s crisis intervention strategy for higher medical education in response to COVID-19 outbreak. Although from the perspective of online teaching practice, it is hard for teachers to follow the performance of each student in online classes, but this strategy provides opportunities to turn crisis into opportunity in medical education. In the era of globalization and information, actively reflecting on China’s response and teaching strategies amid the COVID-19 pandemic is of great significance to facilitate the in-depth development of medical science and medical education, deepen the teaching concept of “student-centered” and “competency-oriented” medical education, as well as explore teaching approaches to advance the medical education. More importantly, with the wide spread of COVID-19, China’s experience in online higher medical education may serve as a reference to other countries during the pandemic.

## Data Availability

All data included in this study are available upon request by contact with the corresponding author.

